# A Supervised Learning Framework for Stroke Hospitalization Factors Selection Using the Lasso-MIDAS Model

**DOI:** 10.64898/2026.05.15.26353365

**Authors:** Lu Wang, Qiyang Li

**Author notes:** **Compliance with Ethical Standards Ethical Approval** This study was conducted in strict accordance with the relevant national laws and the institutional ethical requirements of the hospital. All procedures involving human participants were reviewed and approved by the Medical Ethics Committee of The Third People’s Hospital of Chengdu (Approval No.: 2024-S-190). **Informed Consent** The study was performed in line with the principles of the Declaration of Helsinki and local legislative regulations. Given that all data used in this study were de-identified and aggregated, with no direct contact established with individual participants, the institutional review board waived the requirement for written informed consent from participants or their legal guardians/next of kin. No additional consent was deemed necessary for this research. **Data Sources and Availability** This study uses a multi-source mixed-frequency dataset (March 1, 2021–June 30, 2024). Daily stroke hospitalization data were provided by The Third People’s Hospital of Chengdu; restrictions apply to these licensed data, which are available from the corresponding author upon reasonable request. Environmental/meteorological data were obtained from AQISTUDY China (https://www.aqistudy.cn/) and NOAA NCEI (https://www.ncei.noaa.gov/). Monthly socioeconomic indicators were extracted from the National Bureau of Statistics of China (https://data.stats.gov.cn/). All data were retrieved from authoritative institutions to ensure scientific rigor.

## Abstract

Stroke, as an acute cerebrovascular disease with significant public health implications, is influenced by a complex interplay of meteorological conditions, air quality, and socioeconomic factors. However, the inherent challenges of mixed-frequency data from diverse sources and high-dimensional variable spaces limit the effectiveness of traditional regression models.

This study develops a Lasso-MIDAS model framework to identify the key multidimensional drivers of stroke admissions. Using this approach, 21 candidate variables encompassing meteorological, environmental, and economic indicators were screened. The empirical results identified 11 core influencing factors. In the meteorological and environmental dimensions, Wind Speed, Carbon Monoxide (CO), and Sulfur Dioxide (SO2) were identified as significant positive drivers, with Temperature Difference also positively correlating with admission risks. Conversely, Nitrogen Dioxide (NO2) exhibited a negative correlation, potentially reflecting behavioral adaptation and exposure reduction during peak pollution periods. In the socioeconomic dimension, the Consumer Price Index (CPI) for Food, Tobacco, and Alcohol emerged as a major risk factor, highlighting the impact of living cost pressures on public health.

The findings demonstrate the superiority of the Lasso-MIDAS model in handling large-scale healthcare data. It effectively addresses the frequency mismatch problem while enhancing the robustness of causal identification through variable shrinkage. These conclusions provide a scientific basis for health authorities to establish early warning systems and optimize public health policy interventions.

## 1. Introduction

Stroke remains a predominant contributor to the global disease burden, and effective risk management is inherently dependent on the accurate identification of triggers for hospital admissions[1]. The etiological complexity of stroke suggests that risk patterns emerge from the interplay between short-term atmospheric stressors and broader socioeconomic conditions[2].While recent advancements in medical informatics provide access to high-dimensional datasets, utilizing these multi-source variables is technically constrained by the discrepancy in their observation frequencies. Specifically, traditional econometric models rely on uniform temporal alignment, a requirement that often necessitates sacrificing the granularity of mixed-frequency information.

In epidemiological time-series analysis, a systematic “aggregation bias” occurs when high-frequency predictors are forcibly aligned with low-frequency outcomes. To maintain temporal consistency, standard practices often aggregate daily meteorological observations into monthly averages, resulting in “information truncation” [3]. For cerebrovascular research, this temporal smoothing is problematic as it masks acute high-frequency triggers—such as sudden temperature fluctuations or pollutant spikes—that act as direct precursors to stroke events. Consequently, single-frequency models fail to internalize the fine-grained dynamics of these environmental stressors, creating a methodological limitation in identifying the specific drivers of stroke hospitalizations within mixed-frequency datasets.

The objective of this research is to identify the core drivers of stroke hospitalizations from a candidate pool of 21 mixed-frequency variables using the Lasso-MIDAS framework. The scientific contributions of this study are summarized as follows:

### (1) High-frequency Signal Preservation via Mixed-frequency Modeling

Unlike conventional studies that rely on single-frequency alignment (e.g., aggregating daily data into monthly averages), this research introduces a mixed-frequency **(MIDAS)** framework to the domain of stroke surveillance. This approach allows for the direct integration of daily environmental stressors and monthly socioeconomic indicators within a single model. By bypassing traditional temporal down-sampling, the model preserves the original granularity of acute triggers—such as sudden thermal fluctuations—ensuring that high-frequency environmental pulses are not smoothed out or lost.

### (2) Joint Optimization of Variable Selection and Estimation

The research integrates the Lasso penalty into the mixed-frequency structure to achieve simultaneous factor identification and parameter estimation. This methodology automatically filters out redundant variables from the high-dimensional indicator set by shrinking their coefficients to zero[4]. This dual-function mechanism provides a parsimonious solution for identifying core drivers while mitigating the risks of over-fitting and collinearity inherent in multi-source datasets.

## 2. Related work

Since Tibshirani[5] first introduced the Least Absolute Shrinkage and Selection Operator (Lasso), this regularization technique has been extensively applied across diverse scientific domains. In finance and economics, Lasso has demonstrated superior predictive accuracy over traditional OLS and probit models in forecasting credit ratings and crude oil yields, particularly when handling large-scale datasets with complex technical indicators [7][8]. Beyond economics, Lasso has become a critical tool in biomedical research for high-dimensional feature selection. For instance, researchers have utilized Lasso to identify influential genetic markers and clinical variables linked to Alzheimer’s disease and schizophrenia, effectively resolving multicollinearity and overfitting issues in clinical datasets[9]. These applications underscore Lasso’s capacity to extract a parsimonious set of significant predictors from redundant information.

The Mixed Data Sampling (MIDAS) framework, introduced by Ghysels[6], provides a robust methodology for integrating data sampled at different frequencies, such as daily and monthly observations. In energy and macroeconomics, MIDAS has been shown to significantly improve forecasting performance for crude oil prices and GDP growth by internalizing high-frequency market signals[10] . To address the “curse of dimensionality” in multi-source datasets, recent studies have integrated regularization techniques into the MIDAS structure. Notably, Marsilli[4] developed the Lasso-MIDAS model, demonstrating its efficacy in identifying key economic drivers from numerous quarterly indicators. Subsequent extensions, such as MIDASSO (integrating Elastic Net) and Group-Lasso U-MIDAS, have further enhanced the precision of risk assessments and default predictions, proving that penalized MIDAS models can effectively manage both frequency mismatch and high-dimensional noise[11][12] .

In summary, while Lasso regression and MIDAS models have been extensively validated in financial and macroeconomic forecasting, their integrated application within the public health domain—specifically for acute cerebrovascular events—remains limited. Existing epidemiological studies often rely on single-frequency data, which may lead to the omission of high-frequency environmental triggers. By bridging this methodological gap, the present study utilizes a Lasso-MIDAS framework to simultaneously manage frequency discrepancies and high-dimensional indicator selection. This approach not only enhances the precision of stroke hospitalization identification but also provides a more robust tool for analyzing the complex interactions between multi-source environmental and socioeconomic drivers.

## 3. Data Description and Preprocessing

### 3.1. Data Sources

This study constructs a multi-source mixed-frequency dataset spanning from March 1, 2021, to June 30, 2024. To ensure the scientific rigor and international comparability of the research, all data were retrieved from authoritative clinical, environmental, and governmental institutions.

The primary dependent variable, daily stroke hospitalizations, was provided by The Third People Hospital of Chengdu. As a prominent Grade A Tertiary hospital and a national stroke center in Southwest China, its standardized electronic health record (EHR) system ensures that the clinical data are both accurate and representative. Environmental and meteorological data were integrated from the Air Quality Online Monitoring and Analysis Platform (https://www.aqistudy.cn/) and the National Centers for Environmental Information (NCEI) (https://www.ncei.noaa.gov/) of the National Oceanic and Atmospheric Administration (NOAA). These platforms provide globally recognized, high-precision atmospheric observations. Finally, monthly socioeconomic indicators were extracted from the National Bureau of Statistics of China (NBS) (https://data.stats.gov.cn/), the most authoritative source for macroeconomic time-series data in China.

### 3.2. Variables Selection

The selection of candidate variables is grounded in established epidemiological and socioeconomic theories to capture the multi-dimensional drivers of stroke hospitalizations. A total of 21 mixed-frequency indicators were incorporated into the Lasso-MIDAS framework, categorized as follows:

#### Dependent Variable

The daily number of stroke admissions serves as the core dependent variable, representing the acute health outcomes under investigation.

#### Environmental and Meteorological Indicators (Daily)

This category includes 11 high-frequency variables, comprising 7 air quality metrics (AQI, *PM*_2.5_, *PM*_10_, *CO, NO*_2_, *SO*_2_, and *O*_3__8*h*) and 4 meteorological factors (mean temperature, temperature difference, wind speed, and sea-level pressure). These variables are selected because pollutants and sudden atmospheric shifts are recognized as acute triggers that can induce systemic inflammation or cardiovascular stress, directly leading to stroke events.

#### Socioeconomic Indicators (Monthly)

To capture the broader social determinants of health, 10 low-frequency variables were included. These encompass 5 specific Consumer Price Indices (CPI for Clothing, Transport/Communication, Overall, Food/Tobacco, and Healthcare) and 5 industrial activity metrics (Real Estate Investment, Commercial Housing Sales Area, Air Conditioner Output, Automobile Output, and Electricity Generation). These factors reflect the population’s living standards and psychosocial stress levels, which influence long-term cerebrovascular vulnerability.

### 3.3. Data Cleaning and Transformation

To ensure the statistical validity of the Lasso-MIDAS model, a rigorous preprocessing pipeline was applied to the raw multi-source dataset. The primary challenge involved addressing missing values across different frequencies. For the high-frequency daily environmental records, we employed linear interpolation to fill occasional gaps in air quality and meteorological time series, ensuring a continuous sequence. Specifically, for certain monthly socioeconomic indicators—such as automobile production and electricity generation, which contained missing values for January and February—linear interpolation was also utilized to maintain the integrity of the low-frequency data stream.

Furthermore, all 21 candidate variables underwent Z-score normalization to eliminate dimensional discrepancies between heterogeneous data sources. This transformation scales each variable to have a mean of zero and a standard deviation of one, providing a critical prerequisite for the Lasso penalty to perform unbiased variable selection. Finally, we performed temporal alignment to synchronize the daily environmental “shocks” with the monthly socioeconomic “background,” ensuring that the causal structure remains consistent within the mixed-frequency framework.

## 4. Method

### 4.1. Lasso-MIDAS Model

To identify the key drivers of stroke hospitalizations from high-dimensional and mixed-frequency data, this study adopts the Lasso-MIDAS framework. By incorporating an *L*_1_ regularization penalty into the Mixed-Data Sampling (MIDAS) regression, this model enables the automated selection of influential indicators within a unified framework.

The basic specification of the Lasso-MIDAS model is defined as follows:

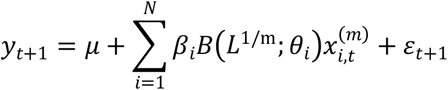

where *y*_*t*+1_ denotes the monthly stroke admissions at time *t* + 1 ; 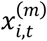 represents the *i*-th daily candidate predictor (including meteorological and air quality indicators) during month *t* ; and *B*(*L*^1/m^; *θ*_*i*_ is the MIDAS polynomial weighting function.

Consistent with the settings in Marsilli[4] and the reference study, to avoid estimation instability of non-linear parameters in a high-dimensional environment, an equal weighting scheme is adopted. Under this specification, the weighting function is simplified as:

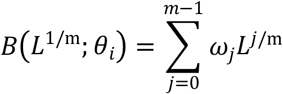

where the weight *ω*_*j*_ = 1/m . This approach effectively compresses the parameter space by aggregating daily shocks within a month into a robust monthly signal, thereby mitigating the “curse of dimensionality” associated with high-frequency lags.

### 4.2. Parameter Estimation and Variable Selection Based on the MAP Algorithm

Traditional estimation methods (such as coordinate descent) often suffer from low computational efficiency when dealing with large-scale variables. To overcome these limitations, this study utilizes the Maximum A Posteriori (MAP) estimation algorithm, as proposed in the reference paper.

#### (1) MAP Estimation Algorithm

The MAP algorithm reformulates the Lasso penalty as a Laplace prior distribution over the parameter vector Γ. By combining the likelihood function with this prior, the parameter estimation is transformed into the maximization of the following posterior density:

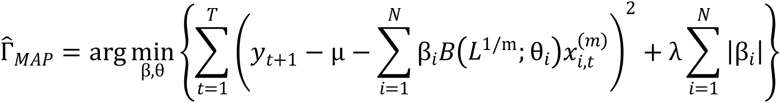

Compared to the original algorithms used in earlier Lasso-MIDAS models, this MAP-based approach offers superior numerical stability and faster convergence speeds, particularly when processing the 21 heterogeneous variables used in this study.

#### (2) Variable Selection and λ Determination

Variable selection is achieved through the shrinkage effect of the *L*_1_ penalty 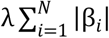. The regularization parameter λ controls the degree of sparsity in the model. As λ increases, coefficients of irrelevant socioeconomic or environmental variables are shrunk toward zero. This study employs **K-fold cross-validation** to identify the optimal λ that balances model parsimony and explanatory power, ensuring that only the most robust drivers of stroke admissions are retained in the final specification.

### 4.3. Evaluation of Model Performance and Explanatory Power

To ensure the reliability of the identified predictors, this study employs core statistical metrics to evaluate the performance of the Lasso-MIDAS model. These metrics assess the consistency between the predicted values 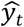 and the actual observations *y*_*t*_:

#### Root Mean Square Error (RMSE)

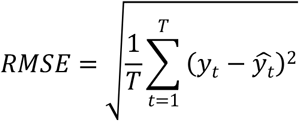

#### Mean Absolute Error (MAE)

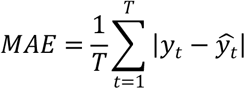

By minimizing these error metrics through the MAP algorithm and cross-validation, we ensure that the selected environmental and socioeconomic factors possess significant and stable explanatory power regarding the fluctuations in stroke hospitalizations.

## 5. Result

### 5.1. Analysis of Variable Selection Results

In this study, the Lasso-MIDAS model was employed to screen factors influencing stroke admissions. By utilizing the *L*_1_ regularization penalty, the model effectively compressed non-significant indicators to zero, identifying 11 key variables from the high-dimensional dataset. The final selection results, which include air quality, meteorological changes, and socioeconomic fluctuations, are presented in Table 1.

**Table 1.**
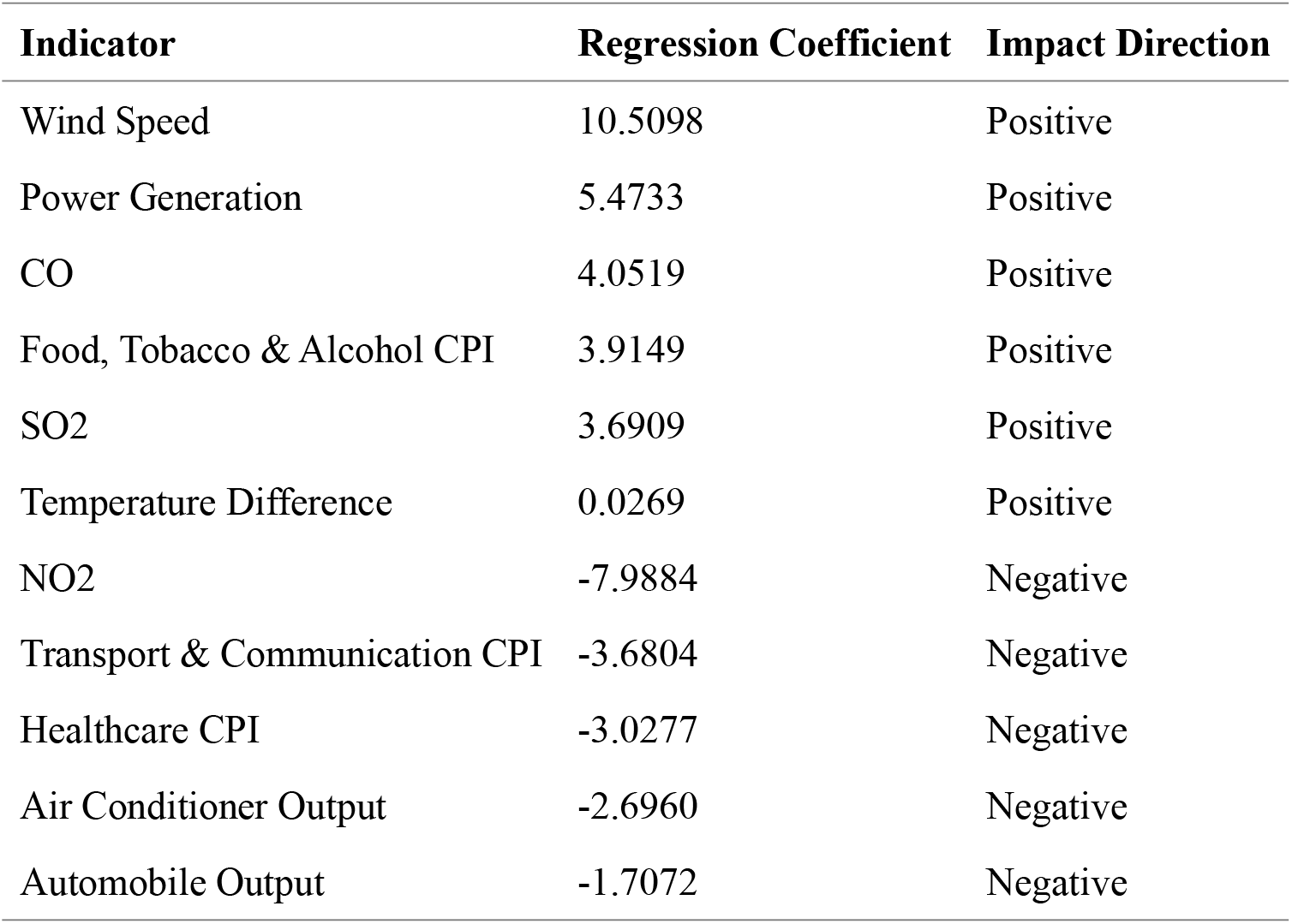
Variable Selection Results Based on Lasso-MIDAS Model.

| Indicator | Regression Coefficient | Impact Direction |
| --- | --- | --- |
| Wind Speed | 10.5098 | Positive |
| Power Generation | 5.4733 | Positive |
| CO | 4.0519 | Positive |
| Food, Tobacco & Alcohol CPI | 3.9149 | Positive |
| SO2 | 3.6909 | Positive |
| Temperature Difference | 0.0269 | Positive |
| NO2 | -7.9884 | Negative |
| Transport & Communication CPI | -3.6804 | Negative |
| Healthcare CPI | -3.0277 | Negative |
| Air Conditioner Output | -2.6960 | Negative |
| Automobile Output | -1.7072 | Negative |

### 5.2. Evaluation of Model Predictive Performance

To verify the fitting accuracy of the Lasso-MIDAS model, Root Mean Square Error (RMSE) and Mean Absolute Error (MAE) were calculated. The relatively low error metrics indicate that the model possesses high goodness-of-fit and robust predictive stability in capturing stroke admission fluctuations.

**Table 2.** Evaluation of Model Fitting Performance.

| Evaluation Metric | Numerical Value |
| --- | --- |
| Root Mean Square Error (RMSE) | 4.0418 |
| Mean Absolute Error (MAE) | 3.3663 |
| Optimal Penalty Parameter (Alpha) | 0.0500 |

## 6. Discussion

### 6.1. Synergistic Effects of Meteorological and Air Quality Factors

The Lasso-MIDAS results identify Wind Speed and CO as primary positive drivers, alongside SO2 and Temperature Difference. The significant positive coefficient of wind speed may seem counterintuitive but often relates to rapid weather system transitions or frontal passages, which create physiological stress. More importantly, the inclusion of CO and SO2—common industrial and traffic pollutants—underscores the direct cardiorenal and vascular toxicity of air pollution. Interestingly, NO2 exhibits a strong negative correlation. In a multi-pollutant model, this may reflect the complex chemical interplay between nitrogen oxides and ozone, or potential behavioral adaptations where populations reduce exposure during specific high-pollution periods.

### 6.2. Socioeconomic Drivers and Public Health Implications

Socioeconomic indicators play a crucial role in the model. The positive impact of Food, Tobacco & Alcohol CPI suggests that rising costs of living and potential increases in unhealthy coping mechanisms (like alcohol or tobacco consumption under stress) correlate with higher stroke risks. Conversely, the negative coefficients for Healthcare CPI and Transport CPI might indicate that as the cost of seeking medical help or transportation increases, non-emergency hospitalizations are delayed, or preventive health behaviors are prioritized. The inclusion of industrial proxies like Power Generation and Automobile Output further captures the influence of macroeconomic cycles and environmental exposure levels tied to industrial activity on regional health outcomes.

## 7. Conclusion

### 7.1. Research Conclusions

This study constructs a Lasso-MIDAS model to identify the multidimensional drivers of stroke admissions. The research findings lead to the following conclusions. High-frequency meteorological and air quality factors significantly influence stroke risks. Wind Speed, CO, and SO2 are identified as major positive predictors, suggesting that atmospheric instability and industrial pollutants serve as critical triggers for acute cerebrovascular events. Furthermore, socioeconomic impact plays a dominant role, with Food, Tobacco & Alcohol CPI influencing long-term admission trends. The selection of industrial proxies such as Power Generation and Automobile Output reflects the indirect pressure of industrial activity and economic cycles on regional public health. Methodologically, the Lasso-MIDAS framework effectively addresses the challenges of mixed-frequency data and high-dimensional variable selection, providing a robust tool for healthcare resource planning.

### 7.2. Policy Recommendations

Local health departments should integrate wind speed and air quality indices into stroke risk forecasting models to issue timely alerts during turbulent weather. Given the impact of living costs on health outcomes, social safety nets should be strengthened during periods of high economic volatility to reduce stress-induced health risks.

### 7.3. Limitations and Outlook

While this study identifies key factors, it is limited by the geographical scope of the data. Future research could incorporate spatial-temporal models to explore the geographical heterogeneity of these drivers across different regions.

## Data Availability

Daily stroke hospitalization data were provided by The Third People's Hospital of Chengdu; restrictions apply to these licensed data, which are available from the corresponding author upon reasonable request. Environmental/meteorological data were obtained from AQISTUDY China (https://www.aqistudy.cn/) and NOAA NCEI (https://www.ncei.noaa.gov/). Monthly socioeconomic indicators were extracted from the National Bureau of Statistics of China (https://data.stats.gov.cn/).

https://www.ncei.noaa.gov/

https://data.stats.gov.cn/

